# While they wait: A cross-sectional survey on wait times for mental health treatment for anxiety and depression for Australian adolescents

**DOI:** 10.1101/2023.08.21.23294348

**Authors:** Mirjana Subotic-Kerry, Thomas Borchard, Belinda Parker, Sophie H. Li, Jayden Choi, Emma V. Long, Philip J Batterham, Alexis E. Whitton, Aniela Gokiert, Lucinda Spencer, Bridianne O’Dea

## Abstract

**Background:** Wait times are reported to impede adolescents’ access to mental health treatment for anxiety and depression. However, there is limited quantitative research on current wait times for the treatment of anxiety and depression for Australian adolescents and the impact of these on young help-seekers.

**Aims:** This study examined Australian adolescents’ experiences of wait times for the treatment of anxiety and depression, including the providers they were waiting to access, the self-reported duration and perceived acceptability of wait times, the association between these wait times and psychological distress, and the support and coping behaviours used by adolescents during this time.

**Method:** From April to June 2022, 375 Australian adolescents aged 13-17 years who were currently waiting, or had previously waited in the past 12 months, for mental health treatment for anxiety and depression completed an anonymous cross-sectional online survey.

**Results:** The mean wait time across all treatment providers was 94.1 days (SD: 69.65). Psychologists and psychiatrists were the most utilised services. Most participants felt their wait times were ‘too long’ and longer wait times were significantly associated with increased psychological distress. Many participants perceived their mental health to have worsened during the wait time and engaged in maladaptive and risky coping behaviours while waiting. Most participants did not receive any support from their healthcare providers during the wait time. However, self-reported treatment attendance remained high.

**Conclusions:** Many Australian adolescents face lengthy wait periods when trying to access mental health treatment and this period may exacerbate distress and maladaptive coping.

## INTRODUCTION

### Wait times for adolescent mental health services in Australia

Anxiety and depression are common mental health problems among adolescents in Australia and worldwide.^1,2^ Although effective treatments exist, long wait times impede access to mental health services and are a major barrier to treatment uptake among youth.^3–5^ Described as the time between initial contact and first appointment,^6^ wait times for adolescent mental health treatment for anxiety and depression continue to rise due to increased demand.^7^ However, wait times for mental health treatment have been found to vary across countries^4,5,8^ and services.^9,10^ In Australia, the lack of transparent reporting on wait times for mental healthcare makes comparisons difficult. Prior to the pandemic, the Australian public youth mental health service headspace reported an average wait of 25.5 days for psychological treatment^3^ and a secret shopper study found a median wait time of 34 days for private psychologists and 41 days for private psychiatrists.^11^ During the COVID-19 pandemic, 88% of surveyed Australian psychologists reported that their wait times had increased, with over half of their clients waiting more than three months for their first session.^12^ While similar patterns of increased demand and long wait times for mental healthcare have been reported in the US, UK, Canada, and other countries,^4,5,8,10^ the current wait times for mental health treatment in Australia and the impacts of these on adolescents are unclear.

### The impact of extended wait times on youth mental health

Evidence is emerging on the potential negative consequences of extended wait times on young people’s mental health and treatment uptake. In general, the wait time between referral and treatment access has been identified as a period of significant vulnerability for adolescents and their families as individuals’ symptoms can be acute, but treatment has not yet begun. Prolonged wait times are associated with the premature termination of treatment,^13^ lower rates of kept appointments,^14^ and increased number of missed appointments.^13,15,16^ Research has also found that longer wait times are associated with symptom deterioration and diminished future help-seeking,^17^ with qualitative reports of increased negative emotional and behavioural consequences and worsened psychological health.^18^ Despite these potential negative impacts, there is a scarcity of quantitative data on wait times for adolescent mental health treatment in Australia.

### Waiting list standards for mental health treatment

In many countries, national waiting list standards for mental health treatment have been introduced to monitor the performance of mental healthcare systems.^19^ In 2016, the National Health Service (NHS) in the UK established wait list targets with 75% of referrals for psychological interventions for anxiety and depression to begin treatment within six weeks, and 95% within 18 weeks.^20,21^ This performance benchmarking was found to significantly reduce wait times, with over 90% of referrals having accessed care within six weeks.^22^ The NHS standards have since been updated to include a four-week wait time target for children and young people.^23^ This is consistent with Norway, where the national wait time target for youth mental healthcare is 35 days.^24^ A key hallmark of high performing mental health systems is the timely accessibility and availability of treatment services.^19^ However, due to the lack of national benchmarking of wait times for mental health services in Australia, the overall wait times experienced by young people and the impacts of these remain unknown.

### Objectives of the current study

The current study aimed to explore young people’s experiences of wait times for mental health treatment for depression and anxiety in Australia. This study examined service utilisation, self-reported wait time duration, and perceived acceptability of wait times among Australian adolescents seeking treatment for depression or anxiety. The associations between self-reported wait times and adolescents’ psychological distress as well as any perceived changes in mental health experienced by young people during their wait time were also examined. Lastly, this study explored the support that young people received during their wait time, the coping behaviours that they used while they awaited care, and their self-reported treatment attendance. Based on past studies, it was hypothesised that treatment-seeking Australian adolescents with depression and anxiety would report an average wait time of at least one month for mental health treatment and services.^3,11^ It was also hypothesised that longer wait times would be associated with greater levels of psychological distress. To our knowledge, this is one of the first studies to examine this aspect of mental healthcare service provision among Australian adolescents and will provide much needed insight on how to better support young people as they await care.

## METHOD

### Design

An online cross-sectional survey was administered between April and June 2022. The survey was written specifically for this study in consultation with young people, mental health professionals, and researchers (see Supplementary Material for a detailed description of the survey development and Appendix A for the full survey).

### Ethical approval

The authors assert that all procedures contributing to this work comply with the ethical standards of the relevant national and institutional committees on human experimentation and with the Helsinki Declaration of 1975, as revised in 2008. All procedures involving human subjects/patients were approved by the University of New South Wales Human Research Ethics Committee (HC190382).

### Sample size

The target sample size was 383 participants based on a confidence level of 95%, population size of N=97, 500,^1^ and a margin of error of 5%.

### Participants

Adolescents were eligible to participate if they were aged 13-17 years old, living in Australia, currently waiting to attend their first session of mental health treatment, or had previously waited (in the last 12 months) longer than one week to access their first session of mental health treatment with a mental health professional or service for symptoms of anxiety and/or depression. Adolescents were excluded if they were (i) currently waiting for a follow-up treatment session with a mental health professional or service that they had accessed previously, or (ii) currently waiting or previously waited for a treatment session that was unrelated to anxiety or depression.

### Recruitment, procedure, and consent

Participants were recruited via study information published on the Black Dog Institute’s website and circulated through the Institute’s clinical service partners. Paid social media campaigns on Facebook, Twitter, Instagram, and LinkedIn were utilised. All study advertisements provided hyperlinks to the survey. Prior to commencing the survey, participants were presented with the Participant Information sheet and were required to pass screening questions and a 4-item Gillick Competence Test^25^ to confirm eligibility and their capacity to provide informed consent. Participants who did not answer all the Gillick Competence items correctly were ineligible to participate. All eligible participants provided consent via an online form and all participants who completed the survey received a 20AUD voucher sent via email.

### Survey measures

#### Demographics

Participants were asked to report their age, gender identity, whether they identified as Aboriginal and/or Torres Strait Islander, whether they identified as Lesbian, Gay, Bisexual, Trans, Queer, Intersex, Asexual, or another diverse sexual identity (LGBTQIA+), the Australian State or Territory and postcode they were currently living in, and their educational/employment status. Postcodes were then classified as ‘metropolitan’ or ‘non-metropolitan’ according to the Australian Bureau of Statistics 2016 Australian Statistical Geography Standard.^26^

#### History of mental health

Participants were asked whether they had ever been formally diagnosed with depression and/or anxiety by a health professional and whether they were currently taking medication prescribed by a health professional for depression and/or anxiety.

#### Treatment providers, wait time duration, perceived acceptability of wait time

Participants were asked to review a list of 11 mental health treatment providers and indicate which professionals and services they were currently waiting to see for the first time (i.e., professionals and services they had been referred to, contacted, and made an appointment with). For each of the treatment providers endorsed, participants were asked to report who referred them, the length of time waited between their first contact and attending their first session (how many months, weeks, days, or I don’t know/I can’t remember), and their perception of the wait time (‘too long’, ‘just right/acceptable’, or ‘unsure/I don’t know’).

#### Psychological distress

Psychological distress was measured by the five-item Distress Questionnaire-5 (DQ5).^27^ Participants were asked to indicate the frequency with which they had experienced various thoughts, feelings, and behaviours in the past 30 days from ‘never’ (1) to ‘always’ (5). Total scores range from 5 to 25 with higher scores indicating greater psychological distress, and a threshold of ≥14 as the clinical cut-off. This scale has demonstrated high internal consistency and convergent validity,^27,28^ and has been used in adolescents.^29^ In the current study, the Cronbach’s alpha for the DQ5 was α=.77.

#### Perceived changes in mental health during the wait time

Participants were asked to rate whether their feelings of sadness or worry had improved or worsened during their wait time using a 5-point Likert scale ranging from ‘worse’ (1) to ‘no change’ (3) to ‘better’ (5). Participants also had the option to select ‘does not apply to me’.

#### Support from healthcare providers during the wait time

Using a 5-point Likert scale ranging from ‘not at all important’ (1) to ‘extremely important’ (5), participants were asked to rate how important it was that their healthcare providers helped them manage their depression and anxiety while they awaited their first treatment session. Participants were then asked to rate how supported they felt by their healthcare providers while they awaited treatment using a 5-point Likert scale ranging from ‘not at all supported’ (1) to ‘extremely supported’ (5). Participants were then asked to report whether they had received any of the commonly provided resources during their wait time (e.g., follow-up session or phone call with a GP, contact from the referred professional, information brochures on mental health, and other support services). Two free response questions were asked: “Is there anything that your healthcare providers could have done to better support you during the wait time?” and “What do you think would have helped you the most during your wait time?”.

#### Sources of personal support during the wait time

Participants were provided with a list of 17 sources of personal support and asked to rate how helpful each source was for them during the wait time. Responses were given using a 5-point Likert scale ranging from ‘not at all helpful (1)’ to ‘extremely helpful (5)’, with an additional option of ‘I didn’t seek/receive help from this source’. Participants were able to indicate other sources of support in a free response option.

#### Importance of additional support for parents/guardians during the wait time

Using a 5-point Likert scale ranging from ‘not at all’ (1) to ‘extremely’ (5), participants were asked to rate how important it was that their parents/guardians be provided with some sort of support to help their parents/guardians to cope better during the wait time.

#### Coping behaviours used during the wait time

Participants were asked to select from a list of 26 randomly displayed behaviours that they had used to cope during their wait time. Participants could select all that applied. For analysis, each behaviour was categorised into one of four types: maladaptive, risky, help-seeking, adaptive. A free response option was also provided so that participants could report any coping behaviours that were not listed.

#### Attendance at first session of mental treatment

Participants who were currently waiting to access mental health treatment were asked how likely they were to attend their first session of treatment using a 5-point Likert scale ranging from ‘extremely unlikely’ (1) to ‘extremely likely’ (5). Participants who selected unlikely or extremely unlikely were then provided with a list of 11 reasons for non-attendance and were asked to select all that applied. Participants who had previously waited in the past 12 months to access mental health treatment were asked whether they attended their first session (‘yes’, ‘no’). Participants who reported that they did not attend were also provided with the same list of reasons for non-attendance and asked to select all that applied.

### Data analyses

Data were collected using Qualtrics and then exported to SPSS version 28.0^30^ for analysis. See Supplementary Material for a detailed description of data cleaning processes. Fraudulent and duplicate responses were detected by comparing participants’ details (e.g., email, postcode), IP addresses, patterns and content of survey responses and participants who completed the survey faster than 40% of the average completion time for the entire sample were removed as recommended by Cobanoglu et al.^31^ To determine wait time durations for treatment, the total mean days waited for each professional or service was calculated using the formula Total Months*30.437 + Total Weeks*7 + Total Days waited. Outliers were identified and removed if the reported total days waited exceeded two and a half years. A total of four outliers were removed from the wait time analysis using these criteria.

Differences in wait times between metropolitan and regional/rural areas were examined using Mann-Whitney U tests. To compare wait times against the NHS benchmarks, the total days waited were collapsed into three categories: within 6 weeks (0 to 42 days), within 18 weeks (0 to 126 days), and greater than 18 weeks (127+ days). To determine the association between wait times and psychological distress (DQ-5), zero-order correlations were conducted for those currently waiting only. Free response options were examined using principles of thematic analysis. Two independent raters (TB and EL) reviewed each response to identify common themes and any disagreements were resolved by a third rater (MS-K).

## RESULTS

### Participants

Figure 1 outlines the participant recruitment and study flow. A total of 780 respondents were assessed for study eligibility. The final sample consisted of 375 full completers (64.0% female, mean age: 16.04 years, SD=1.07, range: 13-17). A total of 43.7% of the final sample (*n*=164/375) were currently waiting for their first session of mental treatment and 56.3% (*n*=211/375) had previously waited, in the past 12 months, longer than one week to access their first treatment session. Over half of the sample identified as being LGBTQIA+ (*n*=207/375; 55.2%). The majority lived in metropolitan areas (*n*=264/375; 70.4%) and were secondary school students (*n*=318/375; 84.8%). More than three-quarters of participants had received a formal diagnosis of depression and/or anxiety from a health professional (*n*=292/375; 77.9%) and 46.7% (*n*=175/375) were taking prescribed medication for their mental health.

**Figure 1.**
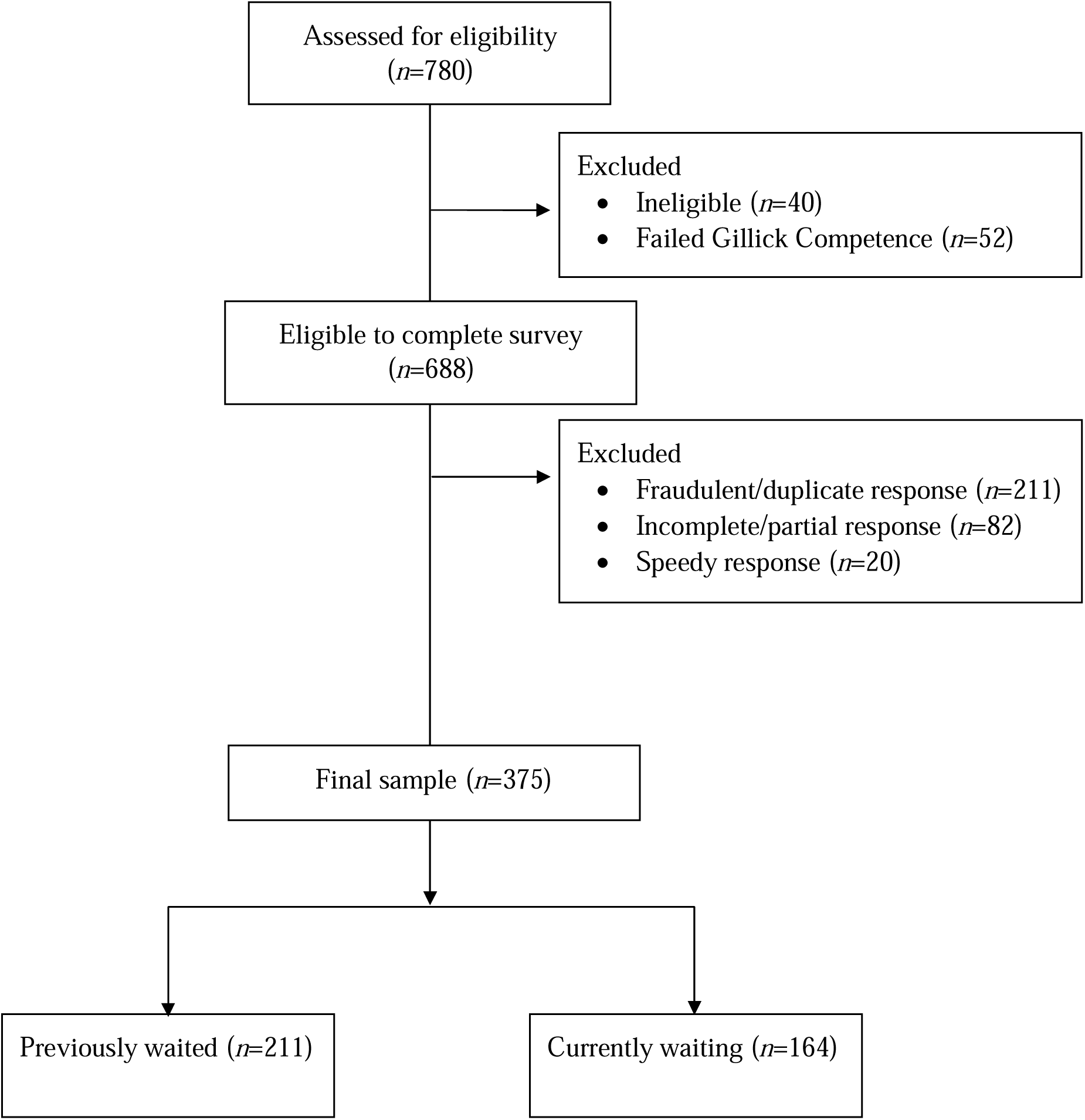
Participant recruitment and study flow diagram.

### Treatment providers, wait time duration and perceived acceptability of wait times

Participants utilised an average of 2.29 (SD: 1.31, range: 1-9) treatment providers. As outlined in Table 2, psychologists (*n*=272; 72.5%) and psychiatrists (*n*=160; 42.7%) were the most common treatment providers. Most participants accessing these were referred by a GP. The mean wait time across all treatment providers was 94.1 days (*SD*: 69.65, range: 5-487, Mdn: 83.85), and the average wait times for the most common treatment providers all exceeded three months. There was significant variability in wait times as demonstrated by the standard deviation estimates ranging from less than one month (21.5 days) to more than two years (744 days). The wait time to access a psychiatrist was significantly longer for those in metropolitan areas compared to regional areas (*U*=1225, *P=*.008). All other comparisons by location did not reach significance (*P= .*082-.943). Across all treatment providers, most participants perceived that their wait time was “too long”.

**Table 1.** Participant demographics (N=375)

|  | N | % |
| --- | --- | --- |
| Gender |  |  |
| Male | 67 | 17.9 |
| Female | 240 | 64.0 |
| Non-Binary | 51 | 13.6 |
| Different Identity | 14 | 3.7 |
| I'd rather not say | 3 | 0.8 |
| Identified as Aboriginal/Torres Strait Islander peoples |  |  |
| Aboriginal peoples | 31 | 8.3 |
| Torres Strait Islander peoples | 1 | 0.3 |
| Aboriginal and Torres Strait Islander peoples | 1 | 0.3 |
| Identified as LGBTQIA+ | 207 | 55.2 |
| Metropolitan location<sup>a</sup> | 264 | 70.4 |
| State or territory of residence |  |  |
| Australian Capital Territory | 5 | 1.3 |
| New South Wales | 107 | 28.5 |
| Victoria | 100 | 26.7 |
| Queensland | 82 | 21.9 |
| Tasmania | 22 | 5.9 |
| Northern Territory | 3 | 0.8 |
| South Australia | 29 | 7.7 |
| Western Australia | 27 | 7.2 |
| Current education or employment status |  |  |
| Secondary school | 318 | 84.8 |
| University | 16 | 4.3 |
| Apprenticeship/Trade/Full-time employment | 12 | 3.2 |
| Other | 29 | 7.7 |
| Formal diagnosis of depression and/or anxiety | 292 | 77.9 |
| Prescribed medication use for depression and/or anxiety | 175 | 46.7 |

*Note.* LGBTQIA+ = Lesbian, Gay, Bisexual, Transgender, Queer, Intersex, and Asexual.

**Table 2.** Treatment providers, wait time durations, and perceived acceptability of wait times among participants (N=375)

| Treatment providers | *n* (%)<br>utilising this<br>service | GP<br>referred<br>*n* (%) | *n* who<br>reported<br>wait time | Mean days waited<br>(SD) | Median<br>days<br>waited | Range<br>(days) | *n* (%) who<br>reported wait<br>time was too<br>long | *n* (%) who<br>reported wait<br>time was<br>acceptable |
| --- | --- | --- | --- | --- | --- | --- | --- | --- |
| Psychologist | 272 (72.5) | 177 (65.1) | 235 | 104.62 (88.5) | 91.3 | 7-574 | 235 (86.4) | 14 (5.1) |
| Psychiatrist | 160 (42.7) | 128 (80.0) | 136 | 149.46 (125.25) | 124.0 | 5-744 | 114 (87.7) | 8 (5.0) |
| School counsellor | 105 (28.0) | 12 (11.4) | 89 | 62.49 (112.44) | 21.0 | 0-727 | 63 (60.0) | 32 (30.5) |
| Headspace | 97 (25.9) | 40 (41.2) | 82 | 103.88 (89.89) | 61.4 | 1-365 | 84 (86.6) | 4 (4.1) |
| Child and<br>Adolescent Mental<br>Health Services | 69 (18.4) | 30 (43.5) | 57 | 77.47 (109.2) | 30.4 | 6-730 | 51 (73.9) | 11 (15.9) |
| Paediatrician | 50 (13.3) | 37 (74.0) | 38 | 167.53 (172.7) | 113.53 | 7-730 | 38 (76.0) | 7 (14.0) |
| Inpatient hospital<br>stay | 32 (8.5) | 17 (53.1) | 27 | 58.9 (69.13) | 30.4 | 1-272 | 22 (68.8) | 4 (12.5) |
| Support group | 27 (7.2) | 6 (22.2) | 18 | 72.02 (78.85) | 43.2 | 14-304 | 14 (51.9) | 8 (29.6) |
| Structured<br>psychological<br>program or service | 25 (6.7) | 9 (36.0) | 19 | 107.94 (130.92) | 83.87 | 3-548 | 13 (52.0) | 7 (28.0) |
| Aboriginal/Torres<br>Strait Islander<br>medical centre | 4 (1.1) | 3 (75.0) | 2 | 45.66 (21.52) | 45.66 | 30-61 | 4 (100.0) | 0 (0) |

### Comparisons with NHS benchmarks

Table 3 outlines the proportion of participants who accessed their first treatment session within the NHS benchmarks. Averaged across all primary health service providers (psychologist, Headspace, psychiatrist, Child and Adolescent Mental Health services), only 28.5% of participants reported a wait time of less than 6 weeks (*n=*146/512). Of these, the proportion that accessed their first treatment session within the 6-week NHS benchmark was lowest for psychiatrists (*n*=21/136; 15.4%), psychologists (*n*=68/235; 28.9%), and headspace centres (*n=*28/84; 33.3%). Just over two-thirds (68.9%) had their first treatment session within 18 weeks and 31.1% waited over 18 weeks.

**Table 3.**
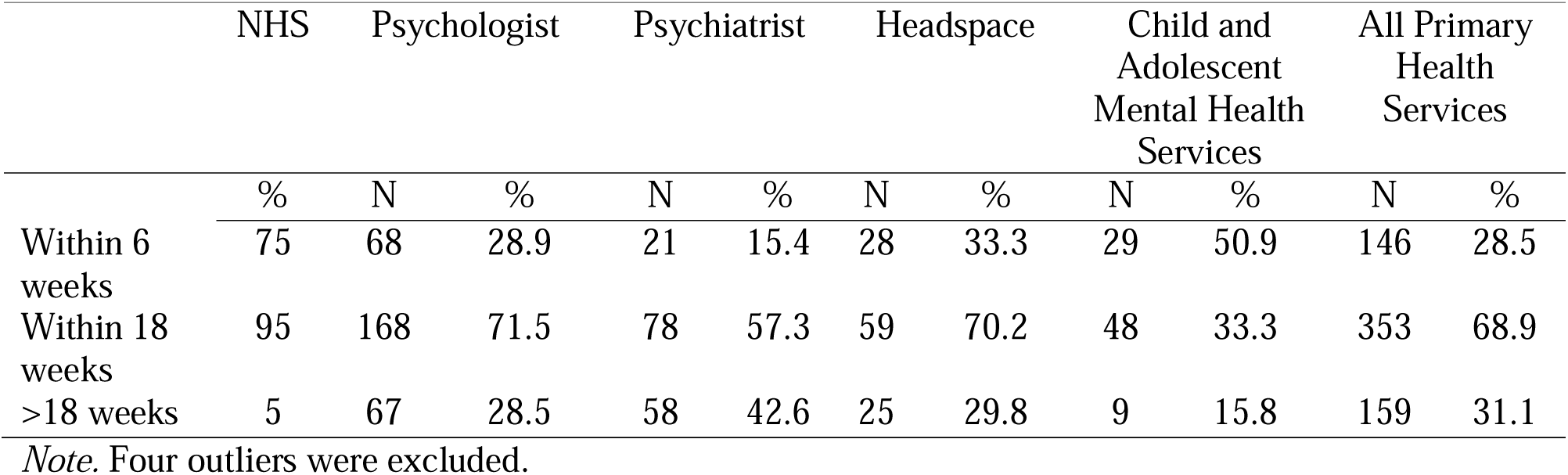
The proportion of participants that received their first treatment session within the NHS benchmarks.

### Psychological distress and perceived changes in mental health during the wait time

Across the whole sample, the mean psychological distress score was 19.40 (*SD*: 3.42, range: 5-25) with 93.3% experiencing clinically meaningful levels of psychological distress. Across the whole sample, 67.5% (*n*=243/360) perceived that their feelings of sadness had worsened during their wait time and 71.5% (*n*=256/363) perceived that their feelings of worry had worsened. In contrast, 13.9% (*n*=50/360) perceived that their feelings of sadness had reduced during their wait time and 14.6% (*n*=53/363) perceived that their worry had reduced.

### Associations between wait times and psychological distress among those currently waiting for their first treatment session

Participants who were currently waiting for their first treatment session reported a mean psychological distress score of 19.13 (*SD*: 3.83, *n=*164) with 90.2% experiencing clinically meaningful levels of psychological distress. In this group, there was a small positive correlation between psychological distress and overall wait times for all services combined (*n*=131, *r*=.23, *P*=.009). There was also a small positive correlation between psychological distress and the wait time for psychologists (*r*=.34, *n*=92, *P*=.001) and psychiatrists (*r*=.31, *n*=43, *P*=.046), such that longer wait times were associated with increased psychological distress. No other significant associations were found (*P*=.117 to .962). Results using Pearson correlations were comparable in magnitude and statistical significance.

### Support from healthcare providers during the wait time

The majority of participants reported that it was ‘very’ or ‘extremely’ important (*n=*274; 73.1%) that their healthcare providers offered them support while they waited for their first treatment session. However, nearly 40% reported that they were ‘not at all’ (*n=*142; 37.9%), or only ‘slightly’ supported (*n=*131; 34.9%) during this time. When asked to select what support they had received, 38.1% (*n*=143) were contacted by their waitlisted provider, 31.2% (*n*=117) had a follow-up session with their GP, 30.9% (*n*=116) were given information on support services, 22.1% (*n*=83) were provided mental health information/brochures, and 21.2% (*n*=79) had received a follow-up phone call from their doctor/GP.

When asked what treatment providers could have done to better support them (free response), the key themes were: increased contact from the waitlisted service (*n*=64/142; 45.1%, e.g., “more check ins”, “greater communication”, and “transparency”), practical information (*n*=48/142; 33.8%, e.g., “mental health strategies and resources” and “online resources”), and other (*n*=30/142; 21.1%, e.g., “crisis support”, “emotional support and validation”, “alternate referrals”, “medication”). When asked what would have helped them the most during the wait time (free response), participants (*n*=71/340; 20.9%) reported “more frequent check-ins” and “greater contact from healthcare providers with updates about the status of appointment”. Participants also requested “resources” (*n*=57/340; 16.8%), “emotional support” or “someone to talk to” (*n*=52/340; 15.3%), “alternate services” or “referral to another mental health professional” (*n*=49/340; 14.4%), “shorter wait times” (*n*=36/340; 10.6%), and support from informal sources such as “parents, friends, and support groups” (*n*=35/340; 10.3%).

### Sources of personal support during the wait time

Table 4 outlines the sources of support participants utilised and associated helpfulness ratings. Most participants turned to friends (*n*=338, 90.1%), parents (*n*=331, 88.3%), and their GP (*n*=305, 81.3%) for support during the wait time. Over half of the sample had used a digital source of support including web-based tools, mental health websites, helplines, and mobile apps. On average, friends were rated as ‘moderately helpful’ sources of support, with all other informal, professional, and digital sources mostly rated as ‘somewhat helpful’. Most participants endorsed that it was ‘very’ to ‘extremely’ important that their parents/guardians be provided with additional support to help them cope during the wait time (*n=*225/375, 60.0%), with very few reporting that it was ‘not at all’ important (*n=*23/375, 6.1%).

**Table 4.** Sources of support used by participants during the wait time (N=375)

| Source of support | Used this source | Helpfulness rating |  |
| --- | --- | --- | --- |
| | $n$ (%) | M | SD |
| Informal sources |  |  |  |
| Friends | 338 (90.1) | 3.09 | 1.18 |
| Parent | 331 (88.3) | 2.30 | 1.18 |
| Siblings | 260 (69.3) | 2.00 | 1.13 |
| Other relative/family | 225 (60.0) | 1.97 | 1.20 |
| Other adult | 201 (53.6) | 2.16 | 1.15 |
| Professional sources |  |  |  |
| GP/local doctor | 305 (81.3) | 2.23 | 1.10 |
| School counsellor | 278 (74.1) | 2.17 | 1.22 |
| Teacher | 257 (73.3) | 2.06 | 1.13 |
| Year advisor or equivalent | 233 (62.1) | 1.94 | 1.15 |
| Other MH professionals | 232 (61.9) | 2.35 | 1.21 |
| Digital sources |  |  |  |
| Web-based assessment tools | 274 (73.3) | 2.56 | 1.18 |
| Mental health websites | 270 (72.0) | 2.40 | 1.21 |
| Telephone helpline | 230 (61.3) | 1.93 | 1.17 |
| Mental health mobile app | 214 (57.1) | 2.00 | 1.00 |
| Online mental health program | 196 (52.2) | 2.06 | 1.10 |
| Online mental health chat services | 189 (50.4) | 2.10 | 1.10 |
| Online mental health support forums | 165 (44.0) | 2.25 | 1.31 |

*Note.* Percentages are reported for the subset of participants that selected each source of support. The range for each source of support listed is 1 (not at all helpful) to 5 (extremely helpful).

### Coping behaviours used during the wait time

As outlined in Table 5, 92.8% (*n*=348) of participants used one or more maladaptive coping behaviours during the wait time such as spending more time alone (*n*=270; 72.0%) and sleeping (*n*=260; 69.3%). A total of 87.5% (*n*=328) used one or more help-seeking behaviours such as searching the Internet to find mental health information (*n*=240; 64.0%) and reaching out to friends via SMS (*n*=199; 53.1%). Over two thirds reported that they had engaged in one or more risky coping behaviours (*n*=284, 75.7%) such as self-harm (*n*=209; 55.7%) and skipping school (*n*=174; 46.4%).

**Table 5.** Coping behaviours used by participants during the wait time (N=375)

|  | *n* | % |
| --- | --- | --- |
| Maladaptive behaviours | 348 | 92.8 |
| Spending more time by myself | 270 | 72.0 |
| Spending more time sleeping | 260 | 69.3 |
| Spending more time on social media | 244 | 65.1 |
| Spending more time at home | 244 | 65.1 |
| Eating more treat food and/or takeaway food | 176 | 46.9 |
| Spending more time online gaming | 106 | 28.3 |
| Help-seeking behaviours | 328 | 87.5 |
| Searching the internet for information about mental health | 240 | 64.0 |
| Speaking with friends over text message | 199 | 53.1 |
| Seeking support from friends | 166 | 44.3 |
| Speaking with a school counsellor, teacher, or other school support | 120 | 32.0 |
| Speaking with friends over a phone call | 111 | 29.6 |
| Risky behaviours | 284 | 75.7 |
| Self-harming | 209 | 55.7 |
| Skipping school | 174 | 46.4 |
| Drinking alcohol | 102 | 27.2 |
| Vaping | 86 | 22.9 |
| Using cannabis | 66 | 17.6 |
| Smoking cigarettes | 49 | 13.1 |
| Using other drugs | 40 | 10.7 |
| Adaptive behaviours | 272 | 72.5 |
| Writing down how I feel (e.g., journaling) | 116 | 30.9 |
| Doing more exercise or sport | 112 | 29.9 |
| Doing activities that help me relax | 111 | 29.6 |
| Reading books | 100 | 26.7 |
| Doing more activities I enjoy | 98 | 26.1 |
| Taking up a new activity, sport, or hobby | 90 | 24.0 |
| Meeting up with friends or becoming more social | 88 | 23.5 |
| Improving or changing my diet | 87 | 23.2 |

*Note.* Total *n* and % for each category were calculated based on whether participants
endorsed at least one strategy in that category.

### Self-reported attendance at the first treatment session

Among those who were currently waiting, 78.7% (*n=*129/164) reported that they were likely to attend their first treatment session and 14.7% (*n=*24/164) reported that they were unlikely to attend. The most common reasons for likely non-attendance were ‘the wait time was too long’ (*n=*13/24; 54.2%), ‘don’t want to go’ (*n=*13/24; 54.2%), and ‘couldn’t be bothered’ (*n=*11/24; 45.8%). Four participants in this subgroup (*n=*4/24; 16.6%) selected the response ‘I don’t need it anymore, I feel better’. Among those who had previously waited, almost all reported that they attended their first session (*n=*203/211; 96.2%); however, ‘the wait time was too long’ (*n=*6/8; 75%) and ‘didn’t want to go’ (*n=*3/8; 37.5%) were the main reasons for self-reported non-attendance in this subgroup.

## DISCUSSION

### Primary findings

This study presents a cross-sectional examination of Australian adolescents’ experiences of wait times for mental health treatment for anxiety and depression. Consistent with the hypotheses, the average self-reported wait times for several mental health treatment providers exceeded 100 days. Most young people in this sample were waiting to access psychologists, psychiatrists, and headspace centres for more than three months and the majority felt that their wait times were ‘too long’. While there was significant variation in wait times across services and between participants, these did not differ between states, and metropolitan location was found to only be significantly associated with greater access to a psychiatrist. The average self-reported wait times found in this study were more than three times higher than previous Australian reports,^3^ although consistent with more recent data on psychologist wait times.^12^ Overall, these results indicate significant gaps between adolescents’ need for mental health treatment for anxiety and depression and its timely availability in Australia.

In further support of our hypotheses, longer wait times were associated with higher levels of psychological distress, and over two-thirds of participants felt their mental health had worsened during the wait time. Moreover, many of the maladaptive and risky coping behaviours used by participants may have signified further deterioration of symptoms (e.g., sleeping, social withdrawal, self-harm). While some participants felt their mental health had improved during the wait time, our results are consistent with several past studies that observed declines in mental health among young people waiting for care.^32–35^ However, as our study is cross-sectional, there was no evidence to suggest that wait times caused poorer mental health in young people. Rather, our results may simply reflect the natural illness progression of anxiety and depression among this sample and their greater need for treatment. Regardless however, our findings suggest that the wait time for mental health treatment is likely to be a period of significant vulnerability for many adolescents, characterised by high levels of psychological distress, perceived worsening of mental health, and engagement in maladaptive and risky coping behaviours.

### Implications for clinical practice

This study confirms that many participants were provided with nil to minimal support from their healthcare providers during the wait time, despite the majority feeling that it was important. Interestingly, the support preferences of participants were low intensive, non-clinical, and communication-based. Specifically, young people requested more contact and ‘check-ins’ from their waitlisted service provider, which could be administered by practice staff or automated through technological platforms such as SMS. As young people endorsed the helpfulness of some digital resources, a system that contacts young people periodically about their appointment, provides links to web-based tools and information, as well as positive coping behaviours, is likely to be regarded as helpful to adolescents on wait lists for anxiety and depression treatment. Future research should actively engage with young treatment seekers to co-develop such an approach. Moreover, the high referral rates and interim care provided by GPs further confirm the importance of their role in mental health service provision in Australia. Future research would benefit from examining GPs’ understanding of wait times, the impacts on their treating behaviour, and how to best support GPs in providing interim care to their youth patients on wait lists for mental health treatment.

In this study, most participants reported that they attended their first treatment session or were likely to, despite experiencing long wait times. This finding contrasts with several studies that imply longer wait times lead to treatment disengagement across adolescents.^13–16^ Our results may reflect the ‘sunken cost’ associated with longer wait times, such that the time, effort, and resources involved in accessing scarce treatment lead to higher levels of retention in youth. This finding may also reflect the higher levels of motivation and commitment to treatment among this sample, which may or may not be due to longer wait times. As most participants were in secondary school, their treatment adherence may have also been sustained through parental, familial, and school support. As such, different patterns of service use may be found in other samples and studies with longer periods of observation. However, long wait times were reported as the primary reason that non-attenders did not start their treatment. This suggests that long wait times may reduce treatment uptake in a sub-group of adolescent help-seekers and future research may benefit from examining this pattern of treatment engagement in more detail. Moreover, international studies have found that many parents facing long wait times place their adolescent children on multiple wait lists, which may further exacerbate wait times.^36,37^ Future studies may benefit from examining whether long wait times lead to over-servicing of treatment providers in Australia.

### The call for national standards

The overall wait times reported in this study exceeded the NHS standards, with only 1 in 4 young people reporting a wait time of less than 6 weeks and one-third waiting longer than 18 weeks. Given that the introduction of transparent wait time standards in the UK and other countries has reduced wait times significantly,^19,22^ our results support the call for transparent wait time monitoring and reporting for mental health treatment in Australia. This approach may improve the timely provision of mental health treatment to both adolescents and adults. As a start, this could be achieved through mandatory reporting from any mental health professional that benefits from the Better Access initiative - a Federal government program that provides subsidised mental healthcare to Australian residents.^38^ This approach would also enable the identification of locations and treatment services with greater need as well as the objective data needed to evaluate the impact of systemic changes on wait time durations.^39^ Future research should utilise evidence-based approaches that involve service users, including clinicians, parents and families, schools, and young people to determine acceptable wait time targets for the Australian context.^40^

### Limitations

This study provides an important step toward assessing wait time data for adolescent mental health treatment for anxiety and depression in the absence of more robust methods of national data collection. A key limitation of the current study relates to the sampling method, such that we may not have captured the views of adolescents who attended their first treatment session within a short timeframe (e.g., less than one week) or who were satisfied with their wait time. Moreover, as well as having a high rate of female participation, over half the sample identified as being LGBTQIA+ which may indicate a sampling bias or may also reflect the greater need for treatment and higher rates of help-seeking in adolescent females and youth who identify as sexuality diverse.^41,42^ There is emerging evidence in the US that rates of LGBT identification are increasing in younger generations, and women were also more likely to identify as sexuality diverse than men.^43^. Further, a recent study ^44^ examining the acceptability and proximal effects of an open-access platform offering three online single-session interventions for youth internalizing distress, reported a large proportion of females (78.10%) and youth identifying as LGBTQIA+ (50.13%) which are comparable to the rates found in this study. Alternatively, although no formal efforts were made to recruit members of specific groups, our recruitment methods may represent efficient avenues for reaching females and sexuality diverse youth. The self-report data may also be limited by poor or inaccurate recall. Different results may be found in treatment provider records or when more objective measures are used. Seasonal variations in wait times reported by other service providers^3^ were also unable to be captured by this study due to the time-limited and cross-sectional study design. As such, different wait times may be found when data is collected over longer periods of time.

## Conclusion

This study is the first to examine Australian adolescents’ wait times for the treatment of anxiety and depression. Findings indicated that many Australian youth face extended delays across several treatment providers, with many adolescents perceiving the wait times as too long. The findings highlight the need for national transparency and benchmarking of wait times for mental health treatment providers in Australia. Many participants felt unsupported by their referred providers and that their mental health had worsened during the wait time, with many engaging in unhelpful coping behaviours. As such, more research is needed to determine best practices for addressing young people’s mental health needs while they await professional treatment for anxiety and depression.

## Data Availability

All data produced in the present study are available upon reasonable request to the authors

## Acknowledgments

We are grateful to the individuals from the Black Dog Institute Youth Lived Experience group for their time and support in the development of the survey. The authors would also like to thank the young people who took part in this study.

## Author contributions

BOD conceived the project and prepared the initial proposal for the funding application with assistance from TB, BP, SL, AEW, AG, and LS. BOD, BP and TB led the development of the survey. BP, TB, MSK and EL provided research and operational support. MSK, TB, BP, JC, and JC analysed the data with statistical support from PJB. MSK wrote the first draft of the manuscript with all authors providing feedback. All authors reviewed and approved the final manuscript.

## References

1. Lawrence HJ, Johnson SE, Saw S, Buckingham WJ, Sawyer MG, Ainley J, et al. Key findings from the second Australian Child and Adolescent Survey of Mental Health and Wellbeing. Aust NZ J Psychiatry [Internet]. 2015;50(9):876–86. Available from: 10.1177/0004867415617836

2. World Health Organisation. Mental health of adolescents [Internet]. 2021. Available from: https://www.who.int/news-room/fact-sheets/detail/adolescent-mental-health

3. Headspace. Increasing demand in youth mental health: A rising tide of need [Internet]. National Youth Mental Health Foundation. 2019. 14 p. Available from: https://headspace.org.au/assets/Uploads/Increasing-demand-in-youth-mentalh-a-rising-tide-of-need.pdf

4. Kowalewski K, McLennan JD, McGrath PJ. A preliminary investigation of wait times for child and adolescent mental health services in Canada. J Can Acad Child Adolesc Psychiatry. 2011;20(2):112–9.

5. Smith J, Kyle RG, Daniel B, Hubbard G. Patterns of referral and waiting times for specialist Child and Adolescent Mental Health Services. Child Adolesc Ment Health [Internet]. 2018;23(1):41–9. Available from: https://www.apa.org/monitor/2023/04/mental-health-services-wait-times

6. Lewis AK, Harding KE, Snowdon DA, Taylor NF. Reducing wait time from referral to first visit for community outpatient services may contribute to better health outcomes: a systematic review. BMC Health Services Research. 2018;18(1):869.

7. Australian Government Productivity Commission. Report on Government Services 2022: 13 Services for mental health. Impact of COVID-19 on data for the Services for mental health section; 2022. Available from: https://www.pc.gov.au/ongoing/report-on-government-services/2022/health/services-for-mental-health

8. NHS Scotland. Child and Adolescent Mental Health Services waiting times in Scotland. Quarter ending 30 June 2013. Public Health Scotland. 51 p. Available from: https://publichealthscotland.scot/media/8975/2021-09-07-camhs-waiting-times-report.pdf

9. Stringer H. Providers predict longer wait times for mental health services. Here’s who it impacts most: Psychologists worry that patients from marginalized populations, particularly people of color, will suffer most amid a worsening workforce shortage. Monitor on Psychology [Internet]. 2023; 54(3); 28. Available from: https://www.apa.org/monitor/2023/04/mental-health-services-wait-times

10. Edbrooke-Childs J, Deighton J. Problem severity and waiting times for young people accessing mental health services. BJPsych Open [Internet]. 2020;6(6):e118. Available from: 10.1192/bjo.2020.103.

11. Mulraney M, Lee C, Freed G, Sawyer M, Coghill D, Sciberras E, et al. How long and how much? Wait times and costs for initial private child mental health appointments. Journal of Paediatrics and Child Health [Internet]. 2021;57(4):526–32. Available from: 10.1111/jpc.15253

12. Australian Psychological Society. Balancing caseloads and surging demand: Your experience and what we are doing. InPsych [Internet]. 2021; 43(4). Available from: https://psychology.org.au/for-members/publications/inpsych/2021/november-issue-4/balancing-caseloads-and-surging-demand

13. Westin AML, Barksdale CL, Stephan SH. The effect of waiting time on youth engagement to evidence based treatments. Community Mental Health Journal [Internet]. 2014;50(2):221–8. Available from: 10.1007/s10597-012-9585-z

14. Gallucci G, Swartz W, Hackerman F. Brief Reports: Impact of the wait for an initial appointment on the rate of kept appointments at a mental health center. Psychiatric Services [Internet]. 2005;56(3):344–6. Available from: 10.1176/appi.ps.56.3.344

15. Sherman ML, Barnum DD, Buhman-Wiggs A, Nyberg E. Clinical intake of child and adolescent consumers in a rural community mental health center: does wait-time predict attendance? Community Ment Health J [Internet]. 2009;45(1):78–84. Available from: 10.1007/s10597-008-9153-8

16. Williams ME, Latta J, Conversano P. Eliminating the wait for mental health services. J Behav Health Serv Res [Internet]. 2008;35(1):107–14. Available from: 10.1007/s11414-007-9091-1

17. Black G, Roberts RM, Li-Leng T. Depression in rural adolescents: relationships with gender and availability of mental health services. Rural Remote Health. 2012;12:2092.

18. Punton G, Dodd AL, McNeill A. ’You’re on the waiting list’: An interpretive phenomenological analysis of young adults’ experiences of waiting lists within mental health services in the UK. PLoS One [Internet]. 2022;17(3):e0265542. Available from: 10.1371/journal.pone.0265542

19. OECD. A New Benchmark for Mental Health Systems: Tackling the Social and Economic Costs of Mental Ill-Health, OECD Health Policy Studies [Internet]. OECD Publishing, paris. 2021. Available from: 10.1787/4ed890f6-en.

20. Department of Health and Social Care. Achieving Better Access to Mental Health Services by 2020. Department of Health, NHS England. 2014; 23 p.

21. NHS England. Improving Access to Psychological Therapies (IAPT) aiting Times Guidance and FAQ’s. 2015; 16 p. Available from: https://www.england.nhs.uk/wp-content/uploads/2015/02/iapt-wait-times-guid.pdf.

22. NHS 75 Digital. Psychological Therapies: reports on the use of IAPT services, England April 2019 final including reports on the IAPT pilots. 2019. Available from: L https://digital.nhs.uk/data-and-information/publications/statistical/psychological-therapies-report-on-the-use-of-iapt-services/april-2019-final-including-reports-on-the-iapt-pilots.

23. Potter C. NHSE to begin ‘implementation plan’ for new mental health waiting time standards. Pulse 365. 2022.

24. Yang F, Wangen KR, Victor M, Solbakken OA, Holman PA. Referral assessment and patient waiting time decisions in specialized mental healthcare: an exploratory study of early routine collection of PROM (LOVePROM). BMC Health Services Research [Internet]. 2022;22(1):1553. Available from: 10.1186/s12913-022-08877-4

25. Kelly AB, Halford WK. Responses to ethical challenges in conducting research with Australian adolescents. Australian Journal of Psychology [Internet]. 2007;59(1):24–33. Available from: 10.1080/00049530600944358

26. Australian Bureau of Statistics (ABS). 1270.0.55.001 - Australian Statistical Geography Standard (ASGS): Volume 1 - Main Structure and Greater Capital City Statistical Areas, July 2016 [Internet]. 2016. Available from: https://www.abs.gov.au/ausstats/abs@.nsf/mf/1270.0.55.001

27. Batterham PJ, Sunderland M, Carragher N, Calear AL, Mackinnon AJ, Slade T. The Distress Questionnaire-5: Population screener for psychological distress was more accurate than the K6/K10. J Clin Epidemiol [Internet]. 2016;71:35–42. Available from: 10.1016/j.jclinepi.2015.10.005

28. Batterham PJ, Sunderland M, Slade T, Calear AL, Carragher N. Assessing distress in the community: psychometric properties and crosswalk comparison of eight measures of psychological distress. Psychological Medicine [Internet]. 2018;48(8):1316–24. Available from: 10.1017/S0033291717002835

29. Werner-Seidler A, Huckvale K, Larsen ME, Calear AL, Maston K, Johnston L, et al. A trial protocol for the effectiveness of digital interventions for preventing depression in adolescents: The Future Proofing Study. Trials [Internet]. 2020;21(1):2. Available from: 10.1186/s13063-019-3901-7

30. IBM Corp. SPSS Version 28. 2021.

31. Cobanoglu C, Cavusoglu M, Turktarhan G. A beginner’s guide and best practices for using crowdsourcing platforms for survey research: The case of Amazon Mechanical Turk (MTurk). Journal of Global Business Insights. 2021;6(1):92–7.

32. Aisbett DL, Boyd CP, Francis KJ, Newnham K, Newnham K. Understanding barriers to mental health service utilization for adolescents in rural Australia. Rural and Remote Health. 2007;7.

33. Iskra W, Deane FP, Wahlin T, Davis EL. Parental perceptions of barriers to mental health services for young people. Early Interv Psychiatry [Internet]. 2018;12(2):125–34. Available from: 10.1111/eip.12281

34. Leijdesdorff S, Klaassen R, Wairata DJ, Rosema S, van Amelsvoort T, Popma A. Barriers and facilitators on the pathway to mental health care among 12-25 year olds. Int J Qual Stud Health Well-being [Internet]. 2021;16(1):1963110. Available from: 10.1080/17482631.2021.1963110

35. Reichert A, Jacobs R. The impact of waiting time on patient outcomes: Evidence from early intervention in psychosis services in England. Health Economics [Internet]. 2018;27(11):1772–87. Available from: 10.1002/hec.3800

36. Reid GJ, Cunningham CE, Tobon JI, Evans B, Stewart M, Brown JB, et al. Help-seeking for children with mental health problems: parents’ efforts and experiences. Adm Policy Ment Health [Internet]. 2011;38(5):384–97. Available from: 10.1007/s10488-010-0325-9

37. Shanley DC, Reid GJ, Evans B. How parents seek help for children with mental health problems. Adm Policy Ment Health [Internet]. 2008;35(3):135–46. Available from: 10.1007/s10488-006-0107-6

38. Department of Health and Aged Care. Better access to psychiatrists, psychologists and general practitioners through the MBS (Better Access) initiative [Internet]. Australian Government. 2023. Available from: https://www.health.gov.au/our-work/better-access-initiative?utm_source=health.gov.au&utm_medium=callout-auto-custom&utm_campaign=digital_transformation.

39. Rastpour A, McGregor C. Predicting patient wait times by using highly deidentified data in mental health care: enhanced machine learning approach. JMIR Ment Health [Internet]. 2022;9(8):e38428. Available from: https://mental.jmir.org/2022/8/e38428 DOI: 10.2196/38428

40. Eichstedt JA, Singh D, Chen S, Collins KA, Cawthorpe D. Who should be seen when? Establishing wait time benchmarks for children’s mental health. Canadian Journal of Community Mental Health [Internet]. 2021;40(1):105–22. Available from: 10.7870/cjcmh-2021-008

41. Shorey S, Ng ED, Wong CHJ. Global prevalence of depression and elevated depressive symptoms among adolescents: A systematic review and meta-analysis. Br J Clin Psychol [Internet]. 2022;61(2):287–305. Available from: 10.1111/bjc.12333

42. Wilson C, Cariola, LA. LGBTQI+ youth and mental health: A systematic review of qualitative research. Adolescent Research Review, 2020; 5(2), 187–211. 10.1007/s40894-019-00118-w

43. Newport, F. (2018, May 22). In U.S., estimate of LGBT population rises to 4.5 percent. Gallup. https://news.gallup.com/poll/234863/estimate-lgbt-population-rises.aspx

44. Schleider JL, Dobias M, Sung J, Mumper E, Mullarkey MC. Acceptability and Utility of an Open-Access, Online Single-Session Intervention Platform for Adolescent Mental Health. JMIR Ment Health. 2020 Jun 30;7(6):e20513. doi: 10.2196/20513. PMID: 32602846; PMCID: PMC7367540.

