## Supplementary File and Appendix A for "While they wait: A cross-sectional survey on wait times for mental health treatment for anxiety and depression for Australian adolescents"

**Supplementary Material**

1. **Survey Development**

***Step 1 (Expert Consultation)*****:** Once the first survey draft was proposed by the research team, it was reviewed by three academic experts with experience conducting research in the field of adolescent mental health. Each expert evaluated the survey and rated the items on whether they: 1) are essential, 2) should be retained, or 3) should be modified. Where relevant, experts provided free response feedback on each item with reference to their relevance, wording of questions to be appropriate to the target sample, interpretability, and appropriateness of response options. Experts also provided broad advice on each survey section and were asked to indicate whether any concepts were missing. The research team then reviewed the expert evaluation forms and made judgments on whether to remove, retain, or modify each item guided by the expert feedback. Items were retained if more than half (i.e., two) of the experts voted to retain it.

***Step 2 (Youth Lived Experience Group Consultation)*****:** The research team then consulted the Black Dog Institute Youth Lived Experience (YLE) group. This is a diverse group of young people who provide consultation on research projects within the Institute. The research team met with the YLE group to discuss terminology and response options for the survey items, with emphasis on the most appropriate and inclusive way to word questions with respect to our target sample (young people aged 13-17 years old). Following these discussions, the survey was revised by the research team.

***Step 3 (Research Team Consultation)***: The survey was then provided to all members of the research team for final review using the same criteria described in Step 1. The survey was amended based on their feedback and then transcribed to Qualtrics by a paid research assistant.

***Step 4 (Piloting):*** A total of six young people were recruited from the Black Dog Institute Youth Lived Experience group to assess their experience of completing the survey. Eight evaluation questions were included at the end of the survey which provided information on survey difficulty, intelligibility of the instructions, and clarity of the questions and response items from the perspective of the young person. Information was also gathered on the average time it takes to complete the survey in full. Based on this feedback, final adjustments were made prior to the commencement of data collection.

### 2. Management of Fraudulent Respondents and Data Cleaning Processes

***Existing security measures and deterrents:*** Several basic security measures were integrated into the initial survey. The survey platform Qualtrics included the following prevention settings such as fraud detection security measures including bot detection, security scan monitor, RelevantID, and Prevent Indexing and preventing multiple submissions. The Qualtrics platform also includes software to detect IP address locations, thus allowing foreign IP addresses to be blocked. Additionally, the survey included several free-text responses at various points in the survey, making bot completion less likely.

***Fraudulent survey sign-ups:*** Despite the existing security measures, the survey received multiple fraudulent sign-ups between 16$^{th}$ and 18$^{th}$ May 2022, and again on the 31$^{st}$ May 2022. The sign-ups were quickly suspected to be fraudulent due to the large number of responses that came through within a short period compared to the previous recruitment rate. New surveys were completed in quick succession and some survey completions occurred at unlikely times of the day such as early mornings (before 6.00am) or evenings (after 11.00pm). Additionally, these influxes of survey completions did not correspond to an increase in recruitment efforts, specifically, during a period of advertising. An initial review of the survey responses found these responses to be qualitatively different from the survey responses received prior. Considering all these factors, our team suspected that the study had been targeted by fraudulent respondents.

***Response to fraudulent survey respondents:*** In response to the May 2022 attack, we paused recruitment and closed the survey, and contact was made with the University of New South Wales Human Research Ethics Committee (UNSW HREC) on the 18$^{th}$ May, 2022 to provide details of the attack. We spoke with other researchers at the Black Dog Institute who had experienced a similar situation and reviewed the literature for advice on how to manage the situation. A review of all completed surveys was undertaken alongside a review of processes to better understand which factors may have led to the attack. Following these discussions and initial review, we developed a protocol that aimed to 1) enhance measures to prevent future attacks; 2) detail the process for identifying fraudulent or illegitimate respondents; and 3) outline the procedure for managing suspected fraudulent respondents. The protocol was reviewed by the chief investigator of the project and the Trial Steering Committee.

***Prevention of fraudulent survey respondents:*** Following the first attack in May 2022, we added reCAPTCHA software at the beginning of the survey to prevent bot attacks. After consultation with UNSW HREC, we also made changes to the participant information sheet and consent form (PISCF) and the survey instructions to specify “You will only be able to complete the survey once” and “Please note that only one voucher will be issued per participant.” Each voucher was sent manually by a member of the research team after a review and decision was made on the survey data and if a survey respondent was deemed to be a ‘genuine responder’. Several strategies for identifying invalid survey responses were guided by the literature[1] to systematically identify and remove fraudulent or illegitimate respondents. This procedure is outlined below.

***Identifying and withdrawing fraudulent or invalid survey responses:*** After the first attack in May 2022, IP and email addresses were manually checked by two members of the research team. We created a list of criteria in the pattern of survey data that were invalid, inconsistent, and identified as likely to be fraudulent (see Table 1). During the cleaning process, a response was deemed invalid and removed from the dataset based on one or more of the following criteria. First, multiple responses from the same IP or email address (i.e., duplicate responses) indicated that one individual was completing the survey multiple times. Any partial or incomplete survey responses were also removed. Second, invalid postcodes or postcodes that did not match the Australian state or territory reported were flagged as suspicious. A a large number of postcodes from the same area reported by multiple respondents within a short timeframe or block of time was also flagged as suspicious. Third, any participant who completed the survey faster than 40% of the average completion time for the whole sample was flagged as a possible illegitimate or fraudulent responder based on findings that 'speeders' data is significantly different from those above the 40% threshold.[1] Other suspicious activity included the survey responses within a single survey. Specifically, we examined the pattern of survey responses and the content of free-text responses to the questions (see Table 1 for more details). Based on these criteria, two members of the research team (TB and BP) independently reviewed each response and noted whether it should be removed or retained. Any discrepancies were discussed, and the final decision was made by consensus by a separate member of the research team (JC or MSK). Duplicate email and IP addresses and foreign IP addresses were objective indicators of fraud, and these were automatically withdrawn if both independent researchers flagged identical and multiple email or IP addresses. The other individual variables outlined in Table 1 were not enough to identify someone as a potential illegitimate or fraudulent responder; it was the combination of one or more of these characteristics within a single survey completion and similarities between separate survey respondents signing up in short succession or during unlikely times of the day. Further, the two researchers had to be in full agreement regarding the fraudulent or illegitimate categorisation for the respondent to be removed. Data quality checks were conducted regularly to quickly identify suspicious sign-ups and patterns until the survey was closed in June 2022.

Table 1. Indicators of fraudulent activity

| Data Category | Variable | Response characteristic or pattern |
|---|---|---|
| Personal details | IP address | IP address from a country outside of Australia, or a duplicate IP address |
| | Email address | Same email address used |
| | Postcode | Invalid postcode or postcode that did not match the Australian state or territory; Large number of postcodes from the same area (e.g., CBD) within a short timeframe |
| Speed responders | Time spent in survey | Survey completions that take less than 40% of the average time of legitimate respondents |
| Survey responses | Response patterns within a single survey | Survey entries where the respondent has consistently provided the same or similar responses or answered in a pattern for all questions, for example:<br>• All questions answered were "Yes" or all "1's"<br>• All questions answered were "Yes", "No", "Yes", "No" and so forth<br>• Answers in a zig zag (e.g., "1, 2, 3, 4, 3, 2, 1") |
| | Free-text responses | Overuse of Not Applicable. Legitimate free-text responses are predominantly thoughtful and detailed and answer the question being asked. Fraudulent responses mostly use a version of "Not Applicable" in the form of "na", "NA" "none", and this response is often repeated across the free response questions. |
| | | Examples of potential fraudulent responses included:<br>• Using previous matrix question options as answers for future questions<br>• Commonly starting free-text responses in the same way<br>• Duplicate responses across multiple participants<br>• Answers that don't match the question that was asked<br>• Responses indicating that the participant does not live in Australia (e.g., Junior High, Middle School) |

**References**


1. Cobanoglu C, Cavusoglu M, Turktarhan G. A beginner's guide and best practices for using crowdsourcing platforms for survey research: The case of Amazon Mechanical Turk (MTurk). Journal of Global Business Insights. 2021;6(1):92-7.

Appendix A: Survey

| **DEMOGRAPHICS** | |
|---|---|
| 1. What is your age in years? | Numeric response |
| 2. What is your gender identity? | 1. Male<br>2. Female<br>3. Non-binary<br>4. Different identity (please specify)<br>5. I’d rather not say |
| 3. Are you of Aboriginal or Torres Strait Island origin? | 1. Yes – Aboriginal<br>2. Yes - Torres Strait Islander<br>3. Yes - Aboriginal and Torres Strait Islander<br>4. No<br>5. I’d rather not say |
| 4. Do you identify as LGBTQIA+ (Lesbian, Gay, Bisexual, Trans, Queer, Intersex, Asexual, or another diverse sexual identity)? | 1. Yes<br>2. No<br>3. I’d rather not say |
| 5. What state do you currently live in? (If you live in more than one state, please choose the one you spend the most time in). | 1. New South Wales<br>2. Queensland<br>3. Australian Capital Territory<br>4. Victoria<br>5. Northern Territory<br>6. South Australia<br>7. Western Australia<br>8. Tasmania |
| 6. What is the postcode of the suburb where you live? | Numeric response |
| 7. Are you currently… | 1. In high school<br>2. Working full time<br>3. Studying at university<br>4. Completing an apprenticeship<br>5. Other (please specify) |
| **HISTORY OF MENTAL HEALTH** | |
| We would like to hear about your experience accessing mental health treatments and services. In particular, we are interested in learning about “wait times” – the time you waited between contacting your mental health treatment provider or service and your first session.<br><br>Don’t forget, all your answers are anonymous. This means that we have no way of identifying you from your responses. We really appreciate your time and honesty in answering the questions! | |
| 1. Have you ever been formally diagnosed with (or been told that you have) depression and/or anxiety by a health professional (e.g., your doctor/GP, | 1. Yes<br>2. No<br>3. Unsure<br>4. I’d rather not say |

| | |
|---|---|
| psychologist, psychiatrist, school counsellor) | |
| 2. Are you currently taking medication prescribed by a health professional (e.g., your doctor/GP, psychologist, psychiatrist, school counsellor) for depression and/or anxiety (e.g. anti-depressants)? | 1. Yes<br>2. No<br>3. Unsure<br>4. I'd rather not say |
| **EXPERIENCES WHILE WAITING FOR CARE** | |
| **For participants who are <u>currently</u> waiting …** | |
| 1. (A) We would like to hear about the mental health professionals and services that you are currently waiting to see for the first time.<br><br>Please choose the ones you or your parents/guardian, family, or trusted adult have contacted and are now waiting to see.<br><br>Select all that apply. | ☐ Psychologist<br>☐ Psychiatrist<br>☐ headspace centre<br>☐ Hospital stay<br>☐ A program or service to help improve feelings of sadness or worry (e.g. Cool Kids)<br>☐ Local Child and Adolescent Mental Health services (CAMHS) 0/1 School counsellor<br>☐ Paediatrician<br>☐ A support group (e.g. a group of people meeting to share information, experiences, problems and solutions)<br>☐ An Aboriginal/Torres Strait Islander medical centre.<br>☐ An Aboriginal/Torres Strait Islander support worker.<br>☐ Other:(please tell us what it is in the text box). |
| **Q1. (B), (C) and (D) are asked for every treatment and/or service selected in Q1. (A)**<br><br>**"You said you are currently waiting for the following mental health professional or service: [SERVICE]"…** | |
| 1. (B) Who referred you to this service? Was it your…? | 1. Doctor/GP<br>2. School Counsellor/School<br>3. You self-referred (i.e., you or your parents/family booked a session without needing a referral from a doctor)<br>4. Other (please tell us who in the text box)<br>1. I don't know/I can't remember |
| 1. (C) How long will you have waited between contacting this mental health professional or service and going to your first session? | 2. ___Months<br>3. ___Weeks<br>4. ___Days<br>5. ___I don't know/I can't remember |

| | |
|---|---|
| We understand that this can be hard to estimate, so just give it your best go. | |
| 1. (D) Do you think that this wait time is… | 1. Too long<br>2. Just right/acceptable<br>3. Unsure/I don't know |
| 2. Have your feelings of sadness or worry been getting better or worse during your wait time? | Slider from worse to better<br>1 - 2 - 3 - 4 - 5<br>WORSE No Change BETTER<br><br>One slider for sadness One slider for worry<br><br>Option to tick "Does not apply to me" |
| 3. Below is a list of things young people may do to cope while waiting to see a mental health professional or access other services.<br><br>Have you tried any of these?<br><br>Select all that apply. | ☐ Doing more exercise or sport<br>☐ Taking up a new activity, sport, or hobby<br>☐ Improving or changing my diet<br>☐ Seeking support from friends<br>☐ Doing more activities I enjoy<br>☐ Reading books<br>☐ Searching the internet for information about mental health<br>☐ Writing down how I feel (e.g. journaling)<br>☐ Meeting up with friends or becoming more social<br>☐ Speaking with friends over text message<br>☐ Speaking with friends over a phone call<br>☐ Doing activities that help me relax<br>☐ Speaking with a school counsellor, teacher, or other school support<br>☐ Smoking cigarettes<br>☐ Vaping<br>☐ Drinking<br>☐ Using cannabis<br>☐ Using other drugs<br>☐ Self-harming<br>☐ Skipping school<br>☐ Spending more time on social media<br>☐ Spending more time online gaming<br>☐ Eating more treat food and/or takeaway food<br>☐ Spending more time by myself 0/1 Spending more time at home<br>☐ Spending more time sleeping |
| 4. Is there anything else you have been doing to help you cope while you are waiting for your first session? | Free response |

| | |
|---|---|
| 5. Is there anything else you'd like to share with us about how you have been feeling during your wait time? | Free response |
| 6. How likely are you to attend your first session? | 1. Extremely unlikely<br>2. Unlikely<br>3. Neither/Unsure<br>4. Likely<br>5. Extremely likely<br><br>**If select 1 OR 2, go to Q7. Else skip to Q8** |
| 7. Why are you unlikely to attend your first session?<br><br>Select all that apply | ☐ I don't need it anymore because I feel better<br>☐ I found an earlier session somewhere else<br>☐ I have had to wait for too long<br>☐ I can't be bothered<br>☐ I might forget<br>☐ I don't have the money<br>☐ I don't want to go<br>☐ The session is too far away from me<br>☐ I don't have any transport to get there<br>☐ I feel too worried and/or sad to go<br>☐ I am unsatisfied with the service<br>☐ A different reason (please tell us in the text box) |
| 8. How important do you think it is that your healthcare providers (e.g. doctor/GP, psychologists, psychiatrists, school counsellors) help you manage your feelings of sadness and worry while you wait for your first session? | 1. Not at all important<br>2. Slightly Important<br>3. Moderately Important<br>4. Very Important<br>5. Extremely important |
| 9. How supported do you feel by your healthcare providers (e.g. doctors/GPs, psychologists, psychiatrists, school counsellors) while you are currently waiting for your first session? | 1. Not at all supported<br>2. Somewhat supported<br>3. Moderately supported<br>4. Very supported<br>5. Extremely supported |
| 10. Is there anything that your healthcare providers (e.g. doctors/GP, psychologists, psychiatrists, school counsellors) could do to better support you during the wait time? | Free response |
| 11. Overall, what do you think would help you the most during the wait time? | Free response |
| **For participants who have <u>previously</u> waited…** | |
| 12. (A) We would like to hear about the mental health professionals and services that you have accessed for the <u>first time</u> in the past 12 months and have waited <u>more than one week</u> to see. | ☐ Psychologist<br>☐ Psychiatrist<br>☐ headspace centre<br>☐ Hospital stay |

| | |
|---|---|
| Please choose which ones you waited more than one week to see in the past 12 months.<br><br>Select all that apply. | ☐ A program or service to help improve feelings of sadness or worry (e.g. Cool Kids)<br>☐ Local Child and Adolescent Mental Health services (CAMHS)<br>☐ School counsellor<br>☐ Paediatrician<br>☐ A support group (e.g. a group of people meeting to share information, experiences, problems and solutions)<br>☐ An Aboriginal/Torres Strait Islander medical centre.<br>☐ An Aboriginal/Torres Strait Islander support worker.<br>☐ Other:(please tell us what it is in the text box).<br><br>Q9. Displayed as a single question |
| **Q12. (B), (C), and (D) asked for every treatment or service selected in Q12 (A)**<br><br>**"You said you have previously waited for the following mental health professional or service: [SERVICE]"…** | |
| 12. (B) Who referred you to this service? Was it your…? | 1. Doctor/GP<br>2. School Counsellor/School<br>3. You self-referred (i.e., you or your parents/family booked a session without needing a referral from a doctor)<br>4. Other (please tell us who in the text box)<br>5. I don't know/I can't remember |
| 12. (C) From the time you or your family first contacted this service, how long did you have to wait before you had your first actual session?<br><br>We understand that this can be hard to estimate, so just give It your best go.<br><br>How many… | 1. ___Months<br>2. ___Weeks<br>3. ___Days<br>4. ___I don't know/I can't remember |
| 12. (D) Do you think that the wait time was… | 1. Too long<br>2. Just right/acceptable<br>3. Unsure/I don't know |
| 13. Did your feelings of sadness or worry get better or worse while you were waiting? | Slider from worse to better<br>1 - 2 - 3 - 4 - 5<br>WORSE No Change BETTER<br><br>One slider for sadness One slider for worry<br><br>Option to tick "Does not apply to me" |

| | |
|---|---|
| 14. Below is a list of things young people have done to cope while waiting to see a mental health professional or access other services.<br><br>Did you try any of these while you were waiting? Select all that apply. | ☐ Did more exercise or sport<br>☐ Took up a new activity, sport, or hobby<br>☐ Improved/changed my diet<br>☐ Sought support from friends<br>☐ Did more activities I enjoyed<br>☐ Read books<br>☐ Searched the internet for information about mental health<br>☐ Started writing down how I felt (e.g. journaling)<br>☐ Met up with friends or became more social<br>☐ Spoke with friends over text message<br>☐ Spoke with friends over a phone call<br>☐ Did activities that help me relax<br>☐ Spoke with a school counsellor, teacher, or other school support<br>☐ Smoked cigarettes<br>☐ Vaped<br>☐ Drank alcohol<br>☐ Used cannabis<br>☐ Used other drugs<br>☐ Self-harmed<br>☐ Skipped school<br>☐ Spent more time on social media<br>☐ Spent more time online gaming<br>☐ Ate more treat food and/or takeaway food<br>☐ Spent more time by myself<br>☐ Spent more time at home<br>☐ Spent more time sleeping |
| 15. Is there anything else that you did to help cope while you waited for your first session? | Free response |
| 16. Is there anything else you'd like to share with us about how you felt feeling during your wait time? | Free response |
| 17. Did you go to your first session? | 1. Yes<br>2. No<br><br>**If No, go to Q18. If select Yes, skip to Q19** |
| 18. Why didn't you go to your first session?<br><br>Select all that apply. | ☐ I didn't need it anymore because I felt better<br>☐ I found an earlier session somewhere else<br>☐ I had to wait for too long<br>☐ I couldn't be bothered<br>☐ I forgot<br>☐ I didn't have the money<br>☐ I didn't want to go |

| | |
|---|---|
| | ☐ The session was too far away from me<br>☐ I didn't have any transport to get there<br>My parents told me I'm not going<br>☐ I felt too worried and/or sad too go<br>Something came up<br>☐ I was unsatisfied with their service<br>☐ A different reason (please tell us in the text box) |
| 19. How important do you think it was that your healthcare providers (e.g. doctor/GP, psychologists, psychiatrists, school counsellors) helped you manage your feelings of sadness and worry while you waited for your first session? | 1. Not at all important<br>2. Slightly Important<br>3. Moderately Important<br>4. Very Important<br>5. Extremely important |
| 20. How supported did you feel by your healthcare providers (e.g., doctors/GPs, psychologists, psychiatrists, school counsellors) while you waited for your first session? | 1. Not at all supported<br>2. Somewhat supported<br>3. Moderately supported<br>4. Very supported<br>5. Extremely supported |
| 21. Is there anything that your healthcare providers (e.g. doctor/GP, psychologists, psychiatrists, school counsellors) could have done to better support you during the wait time? | Free response |
| 22. Overall, what do you think would have helped you the most during your wait time? | Free response |
| **PARENT SUPPORT** | |
| 1. How important do you think it is that the parents/guardians be given some sort of support to help themselves (parents/guardians) cope better during the wait time? | 1. Not at all important<br>2. Somewhat Important<br>3. Moderately Important<br>4. Very Important<br>5. Extremely important |
| **INTERVENTIONS AND SOURCES OF SUPPORT DURING WAIT TIME** | |
| 1. During the waiting period, did you receive…<br><br>☐ A follow-up session with your doctor/GP?<br>☐ A follow-up phone call from your doctor/GP?<br>☐ Contact from the professional or service you were waiting to see?<br>☐ Information or brochures on mental health from a healthcare provider? | 1. Yes<br>2. No<br>3. I can't remember/I don't know |

| | |
|---|---|
| ☐ Information from a healthcare provider about support services that were available to help you?<br>☐ Other Information or resources (please tell us what in the box) | |
| 2. During your waiting period, did you find the following sources of support helpful for your mental health? Please rate how helpful using a scale of 1 (not at all) to 5 (extremely).<br><br>☐ Parents<br>☐ Siblings<br>☐ Other relative or family member<br>☐ Friends<br>☐ Teacher<br>☐ Year Advisor<br>☐ School Counsellor<br>☐ Other adult (e.g., sports coach, a friend's parent, a person at work)<br>☐ General Practitioner/local doctor<br>☐ Other mental health professional (e.g. psychologist, psychiatrist)<br>☐ Telephone helpline (e.g., Kids Helpline, Lifeline)<br>☐ Websites on mental health (e.g., ReachOut, Beyond Blue)<br>☐ Online self-help mental health program (e.g., programs designed to help improve your symptoms of sadness or worry)?<br>☐ Online assessment tools (e.g., tools that ask you questions and tell you whether you are experiencing anxiety and/or depression)?<br>☐ Online support groups or discussion forums?<br>☐ Online mental health chat services (e.g., eHeadspace)?<br>☐ Mobile app for mental health<br>☐ Someone or something else not listed above (tell us In the box – if there is nothing else, please choose 'I didn't seek/receive help from this source') | 1. Not at all helpful<br>2. Somewhat helpful<br>3. Moderately helpful<br>4. Very helpful<br>5. Extremely helpful<br>6. I didn't seek/receive help from this source |
| **CURRENT MENTAL HEALTH (OPTIONAL)** | |
| We would like to know about how you have been feeling over the last 30 days. We store this information securely and will not share your responses with anyone. You do not have to complete this part of the survey if you don't want to. | |

| | |
|---|---|
| The Distress Questionnaire-5 (DQ-5) The following questions ask about thoughts, feelings, and behaviours that you may have experienced in the last 30 days. Please respond to each question by selecting one box per row.<br><br>In the past 30 days…<br>a) My worries overwhelmed me<br>b) I felt hopeless<br>c) I found social settings upsetting<br>d) I had trouble staying focused on tasks<br>e) Anxiety or fear interfered with my ability to do the things I needed to at work, school, or home | 1. Never<br>2. Rarely<br>3. Sometimes<br>4. Often<br>5. Always |
| **CONCLUSION** | |
| Participants are automatically redirected to a separate survey where they answer the following questions and if provided, their email address are recorded separately from their responses. | |
| 1. Would you like to receive the $20 e-giftcard? This will be sent within 3 business days. | 1. Yes<br>2. No |
| 2. Would you like to receive an email copy of the survey results? | 1. Yes<br>2. No |
| 3. Would you like to hear about other research opportunities related to this project? | 1. Yes<br>2. No |
| 4. Please enter your email address here: | |
| **Thank you for doing the survey. Your responses have been recorded. [end of survey]** | |
